# Clinical case study meets population cohort: Identification of a *BRCA1* pathogenic founder variant in Orcadians

**DOI:** 10.1101/2022.07.18.22276644

**Authors:** Shona M. Kerr, Emma Cowan, Lucija Klaric, Christine Bell, Dawn O’Sullivan, David Buchanan, Joseph J. Grzymski, Regeneron Genetics Center, Cristopher V. van Hout, Gannie Tzoneva, Alan R. Shuldiner, James F. Wilson, Zosia Miedzybrodzka

**Author notes:** These authors contributed equally to this work (Shona M. Kerr, Emma Cowan). Indicates joint senior authors (James F. Wilson, Zosia Miedzybrodzka).

## Abstract

The *BRCA1* pathogenic missense variant c.5207T>C; p.Val1736Ala (V1736A) was multiply ascertained in the clinical investigation of breast and ovarian cancer families from Orkney in the Northern Isles of Scotland, UK. Oral history and birth, marriage and death registrations indicated genealogical linkage of the clinical cases to ancestors from the Isle of Westray, Orkney. Further clinical cases were identified through targeted testing for V1736A in women of Orcadian ancestry attending National Health Service (NHS) genetic clinics for breast and ovarian cancer family risk assessments. Fourteen mutation carriers were identified from predictive tests in relatives of affected members of the index kindred, nine more from NHS diagnostic testing plus a further six obligate carrier females. The variant segregates with female breast and ovarian cancer in clinically ascertained cases. Separately, exome sequence data from 2,090 volunteer participants with three or more Orcadian grandparents, in the ORCADES research cohort, was interrogated to estimate the population prevalence of V1736A in Orcadians. The effects of the variant were assessed using Electronic Health Record (EHR) linkage. Twenty out of 2,090 ORCADES research volunteers (∼1%) carry V1736A, with a common haplotype around the variant. This allele frequency is ∼480-fold higher than in UK Biobank participants. Cost-effectiveness of population screening for a single *BRCA1* founder mutation has been demonstrated at a carrier frequency below the ∼1% observed here. Thus we suggest that Orcadian women should be offered testing for the *BRCA1* V1736A founder mutation, starting with those with ancestry from Westray.

## Introduction

Germline mutation in *BRCA1* confers a high lifetime risk of breast and ovarian cancer^1-3^. Mutation screening for disease-associated variants in *BRCA1* and *BRCA2* is widely available in breast and ovarian cancer. This is not only as an aid to cancer prevention and early diagnosis, but also to guide cancer treatment, e.g. consideration of the use of Olaparib in chemotherapy regimens^4^. Available options for intervention in pre-symptomatic carriers of pathogenic variants in the *BRCA1* gene are surgical (risk-reducing prophylactic bilateral mastectomy, bilateral salpingo-oopherectomy), enhanced screening (e.g. annual magnetic resonance breast imaging) and chemoprevention. Predictive testing of unaffected family members is well established in clinical practice.

In isolate populations, a mutation present in a founding or early member can become widespread in later generations, contributing significantly to the overall disease burden. Pathogenic variants in *BRCA1* and *BRCA2* have been described in several isolate and founder populations worldwide, notably Ashkenazi and Sephardi Jews, and Icelanders^5,6^. Genetic screening programmes focused on founder variants in such genes can be cost-effective^7-10^.

The Orkney Complex Disease Study (ORCADES, part of Viking Genes; www.ed.ac.uk/viking) contains a rich resource of more than 2,000 deeply phenotyped research subjects. All of these participants have at least two grandparents from the Orkney Islands, an archipelago off the North coast of Scotland, UK. More than 90% have three or four such grandparents. There is a high degree of kinship in the ancestral Orcadian population, evidenced both in the extensive genealogies, and in genetic data. Genome-wide analyses of an exhaustive sampling of populations across the British Isles and Ireland demonstrate that Orkney and Shetland are the most divergent and isolated populations of all^11^. Moreover, these Northern Isles have the highest degree of Norse admixture in the British Isles and Ireland, reflecting their unique cultural heritage. This also highlights the potential for a different pool of clinically relevant variants, at different frequencies to mainland Scotland. Research by ourselves and others has demonstrated enrichment of rare and low frequency functional variants in isolated populations, including Orkney^12^. Enriched rare variants of large effect are of particular clinical relevance.

The *BRCA1* pathogenic missense variant c.5207T>C; p.Val1736Ala (V1736A) was multiply ascertained in the clinical investigation of breast and ovarian cancer families from Orkney. We have also generated the exome sequences of 2,090 subjects from the ORCADES study. This research dataset provides an opportunity for analyses of the penetrance of rare clinically relevant variants such as V1736A in the Orcadian population, as well as to derive more accurate and (relatively) unbiased population allele frequencies. Both are increasingly important subjects in clinical genetics, as sequencing of healthy populations becomes more common.

The UK Biobank is a large-scale cosmopolitan biomedical database containing genetic, lifestyle and health information from half a million participants in the UK^13^. Linkage to NHS routine electronic health record (EHR) data adds a longitudinal component to both the ORCADES and UK Biobank cohorts. These research cohorts also allow assessments of the case for population screening for specific genetic variants. Here, we demonstrate that ORCADES offers an exemplar for the use of these analyses at population scale.

## Materials and Methods

### Clinical case note review

In the context of routine assessment of family history of cancer risk, the NHS clinical genetics team responsible for care of patients domiciled in the geographic area of Scotland that includes the Orkney islands recurrently observed the *BRCA1* missense variant, c.5207T>C; p.Val1736Ala, in ovarian and breast cancer patients from Orkney. A pedigree of the “super-kindred” was created, using a combination of oral history from multiple family members and the Scottish Register of Births, Marriages and Deaths. History of cancer in consenting living and deceased family members was confirmed from medical records as part of clinical care of the family.

### Research Volunteer Recruitment and DNA extraction

Recruitment to ORCADES took place from 2005-2011. Eligibility criteria for the volunteer participants were age over 18 years and two or more grandparents born in the Orkney Islands. More than 90% had three or four grandparents from Orkney. The participants attended at least two clinics, one for fasting venepuncture and one for physical measurements, and provided broad-ranging consent for research, including for whole genome sequencing, and for their research data to be linkable to their NHS electronic health records. Blood (or very occasionally saliva) samples from participants were collected, processed and stored using standard operating procedures and managed through a laboratory information management system at the Edinburgh Clinical Research Facility, University of Edinburgh.

### ORCADES Cohort Pedigree Information

Records of the births, marriages and deaths in Orkney are held at the General Register Office for Scotland (New Register House, Edinburgh). These records, along with relationship information obtained from study participants and genealogies available online, were used to assemble a detailed pedigree for ORCADES study participants. This pedigree was constructed using RootsMagic software (S&N Genealogy Supplies), corrected to reflect the genetic kinship between individuals using genotype data. The complete pedigree dates back to ∼900 AD and consists of ∼59,000 individuals.

### Genotyping

DNA from all ORCADES participants was used for genome-wide genotyping on the GSA BeadChip (Illumina) at the Regeneron Genetics Center. Monomorphic genotypes and genotypes with more than 2% of missingness and Hardy-Weinberg equilibrium (HWE) p<10^−6^ were removed, as well as individuals with more than 3% of missingness. Details of genotyping, sample and variant quality control of UK Biobank genotyping data are described in Bycroft *et al* ^13^.

### Exome Sequencing

The “Goldilocks” exome sequence data was prepared at the Regeneron Genetics Center, as detailed for the UK Biobank in Van Hout *et al*.^14^. Briefly, the multiplexed samples were sequenced on the Illumina NovaSeq 6000 platform using S2 flow cells. The raw sequencing data was then processed by automated analysis on the DNAnexus platform^15^ where files were converted to FASTQ format, followed by alignment to GRCh38 genome reference using the BWA-mem^16^. Duplicated reads were identified and flagged using the Picard tool (http://picard.sourceforge.net 2018). Genotypes for each individual sample were called using the WeCall variant caller (https://github.com/Genomicsplc/wecall 2018).

Out of 2,131 sequenced samples, the following were removed: 33 samples that were identified as duplicates, 3 whose genetically-determined sex was discordant with the reported sex, 4 with high rates of heterozygosity/contamination, 2 with low sequence coverage (less than 80% of targeted bases achieving 20X coverage) and 1 discordant with genotyping chip. A PVCF file containing all samples was then created using the GLnexus joint genotyping tool^17^. The “Goldilocks” dataset was created by filtering out genotypes with read depth less than 7 reads, keeping only variants that had at least one heterozygous variant genotype with allele balance ratio greater than or equal to 15% (AB ≥ 0.15), or at least one homozygous variant genotype. This was followed by additional filtering of all variants with more than 10% of missingness and HWE p<10^−6^. A total of 2,090 ORCADES participants (820 male and 1,270 female) passed all exome sequence and genotype quality control thresholds. The variant calls in the ORCADES exome dataset are highly accurate, and the *BRCA1* variant c.5207T>C; p.Val1736Ala rs45553935 segregated correctly in families. Details of the quality control of whole exome sequencing on the 200,000 participants from the UK Biobank are described in Backman *et al* ^18^.

### Verification by Sanger Sequencing

The rs45553935 variant was analysed by targeted Sanger sequencing of PCR products amplified from genomic DNA samples. The primers 5⍰AGCTAAGATCTGAACCCGAGA3⍰ and 5⍰CCACCACGCCCAACTAATTT3⍰ were designed using Primer3 software (Thermo Fisher Scientific) and used to generate a fragment of 586 base pairs for analysis. All 20 heterozygous variant calls from the ORCADES exome dataset were independently verified using this method.

### Haplotype analysis

The ORCADES array genotype data were phased using Shapeit2 v2r837^19^, with the duoHMM option that uses the family-based nature of the data^20^. Prior to phasing, the array genotype data were lifted over from the genome build GRCh38 to GRCh37 using liftOver, followed by quality control against the HRC reference panel with Rayner’s HRC-1000G-check-bim (v4.2.13) script that was downloaded from https://www.well.ox.ac.uk/~wrayner/tools/. Details of phasing of UK Biobank genotyping data have been described^13^. Then, the phased genotypes were used to determine a shared haplotype around rs45553935 using the methods described earlier^21^, all performed using R 4.0.2 (R Core Team 2020 https://www.R-project.org/).

A single variant-based haplotype search was performed to determine the haplotype length between the different ORCADES carrier kindreds, and also with the UK Biobank carrier individuals, using a stepwise approach. Using phased genotype data, starting from the rs45553935 rare variant, one SNP at a time was added to define a haplotype. The procedure was repeated until haplotypes of two individuals (both known carriers) no longer matched, providing variant-level resolution of the haplotype length. The procedure was repeated for all pairs of individuals identified as carriers based on the exome sequencing data, both in the ORCADES and UK Biobank datasets. The shortest shared haplotype from ORCADES was then merged with the shortest shared haplotype from the UK Biobank to compare whether the haplotypes match across the 51 variants shared across genotyping chips. Two megabases of exome sequence around rs45553935 in a carrier from the Healthy Nevada Project^22^ were merged with the corresponding region in a carrier from the ORCADES study. 1974 variants overlapped between the two exomes. A similar approach was then taken to assess potential haplotype sharing. Beginning with the rs45553935 variant, moving one SNP at a time, we compared the two genotypes, repeating the procedure until we came to opposing homozygotes, beyond which haplotypes cannot be shared. Identity-by-descent (IBD) analysis was performed using KING 2.1.5^23^.

### EHR Data Linkage in ORCADES

NHS routine datasets linked to ORCADES participants, including the Scottish Cancer Registry SMR06, were accessed using a secure process as for the Generation Scotland cohort^24^. The ORCADES EHR data release date was July 2021.

## Results

### Pathogenicity and co-segregation with cancer of the V1736A variant

The *BRCA1* c.5207T>C; p.Val1736Ala variant is a conservative amino acid substitution in the carboxyl-terminal domain, a region known to be important in BRCA1 function. *In vitro* studies suggested that the variant disrupts BRCA1 activity^25,26^. Independent evidence for pathogenicity comes from saturation genome editing of *BRCA1* exons in HAP1 cells, which revealed V1736A to be non-functional in cultured cells^27^. A report was published of a severe phenotype patient with ovarian cancer at age 28, short stature, microcephaly and significant toxicity from chemotherapy, with compound heterozygous *BRCA1* variants, *BRCA1*, c.2457delC, and c.5207T>C; as well as loss of heterozygosity (LOH) in associated tumours^28^. This, together with segregation data, led to reclassification of V1736A from a variant of unknown significance to likely pathogenic^28^. The interpretation that V1736A is a pathogenic mutation was corroborated by an expert panel in the Clinvar database^29^, accession VCV000037648, annotated as pathogenic by multiple sources. Our own co-segregation studies in the Orcadian clinical super-kindred (Methods and data available on request) support the pathogenic nature of the mutation, acting as an autosomal dominant.

### Clinical genetics case ascertainment and origin of the variant

As part of clinical practice, nine carrier females were ascertained clinically in diagnostic tests of cancer patients. Fourteen V1736A carriers (eight females and six males) were identified from predictive tests in unaffected relatives of NHS patients. Six female obligate carriers were also identified. This gives a total of 23 positive NHS results in females (see Table 2).

**Table 1.** Frequencies of BRCA1 p.Val1736Ala carriers across a range of genomic datasets

| Dataset | Description | Number of Genomes | Carriers of BRCA1 p.Val1736Ala variant | Cases of HBOC (females) | Cohort Publication Reference |
| --- | --- | --- | --- | --- | --- |
| gnomAD v2.1.1 | Unrelated individuals from genetic studies | 125,748 exomes and 15,708 genomes | 0 | - | 31 |
| Viking Health Study Shetland | Isolate population cohort from Shetland, UK | 2,106 exome sequences | 0 | - | 21 |
| DiscovEHR | Unselected population cohort in Pennsylvania, USA | 92,453 exome sequences | 0 | - | 35 |
| ORCADES | Isolate population cohort from Orkney, UK | 2,090 exome sequences | 20: 13 males and 7 females | 1 | 36 |
| UK Biobank | Cosmopolitan population cohort from UK | 200,643 exome sequences | 4: 1 male and 3 females | 1 | 33 |
| Healthy Nevada | Population health study from Nevada, USA | 26,906 clinical 'Exome+' sequences | 1 (female) | 1 | 22 |
| Breast Cancer Association Consortium (BCAC) | Breast cancer cases and controls, worldwide | 42,671 cases with European ancestry, iCOGS custom genotyping array | 2 | 2 | 34 |
| CanVar-UK | Sequenced exomes of cancer patients England & Wales | 28,936 total probands tested | 9 (females) | 9 | 37 |

**Table 2.**
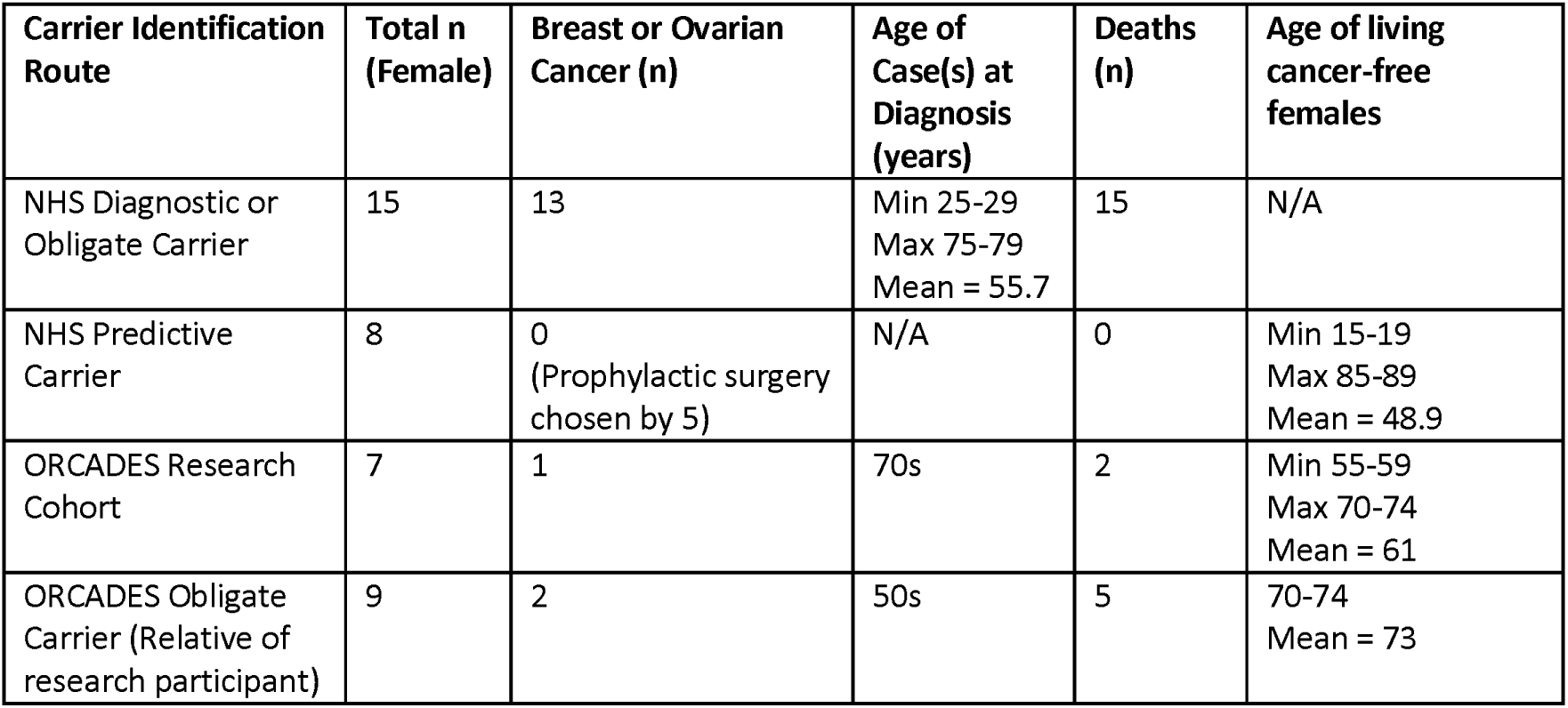
Cancer status of women from Orkney carrying BRCA1 p.Val1736Ala

Analysis of the clinic cases shows the variant segregates in at least two kindreds. Both lines trace back to the 18^th^ century with ancestors from the remote island of Westray, in the North Isles of Orkney.

There are twenty heterozygous carriers of the V1736A variant in the ORCADES exome dataset, of whom seven are female. Consistent with the clinical data, eight of the twenty carriers in ORCADES had all four grandparents born in Westray, and all but one of them had at least one Westray grandparent. Of all 80 grandparents of the carriers, 60% were from Westray, with the majority of the remainder coming from other parishes or isles of Orkney (Figure 1).

**Figure 1.**
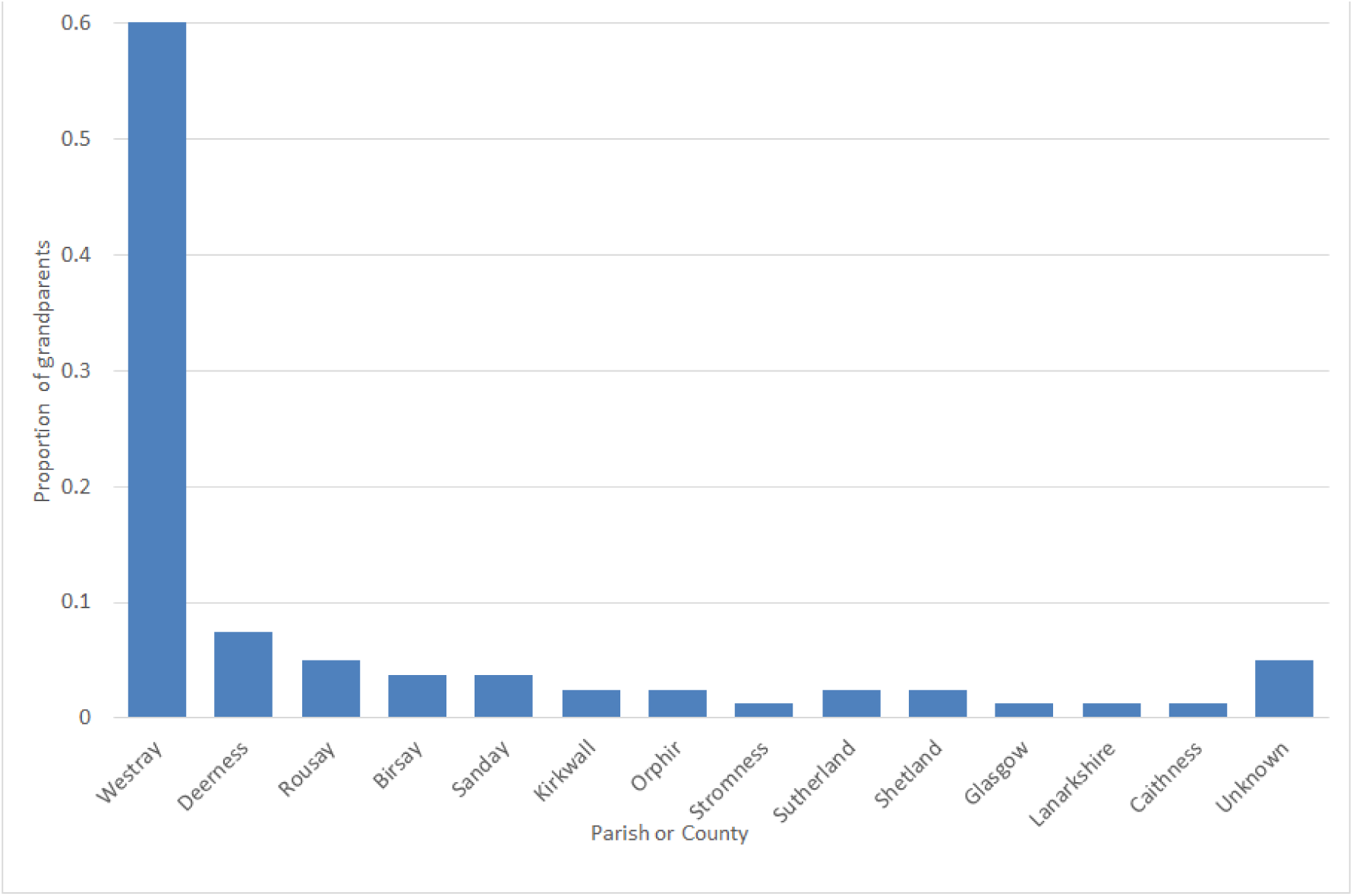
Grandparental ancestry of carriers in ORCADES. The first eight columns are parishes or isles of Orkney. The remainder are locations elsewhere in Scotland, or unknown.

The mutation carriers ascertained clinically and in the ORCADES study can be assembled together into five kindreds descending from five separate couples. There are four kindreds from the ORCADES study and three from the Clinical Genetics dataset, two of which overlap, for the total of five kindreds. While many of these kindreds show distant kinship with one another, e.g. being fifth or sixth cousins, this is not uncommon among individuals with Westray heritage and so it was not possible to be certain of the path of segregation of the variant to each of the carriers living today. Indeed, multiple lines of inheritance were possible in a number of cases. Ancestral non-paternity events may have also influenced the path of segregation down the pedigree. Some of the connections could be prior to *c*.1750, before which paper genealogy records are not generally available. Given the population structure of the island, and limited contribution of ancestors to descendants, it is likely that the kindreds which cannot be linked together genealogically do in fact connect in the preceding few generations at some point before *c*.1700, as demonstrated by their shared haplotype (see below). What is clear is that the ancestry goes back over 250 years in the island of Westray (Figure 2); the founders of kindred A, the largest, having been born there in the 1760s. All four of the other kindreds also eventually lead back to Westray common ancestors, in the 19th century (but with deeper ancestry there back to the same time depth). In kindred C, mostly resident in the East Mainland of Orkney, the Westray common ancestors were born in the early 1800s.

**Figure 2.**
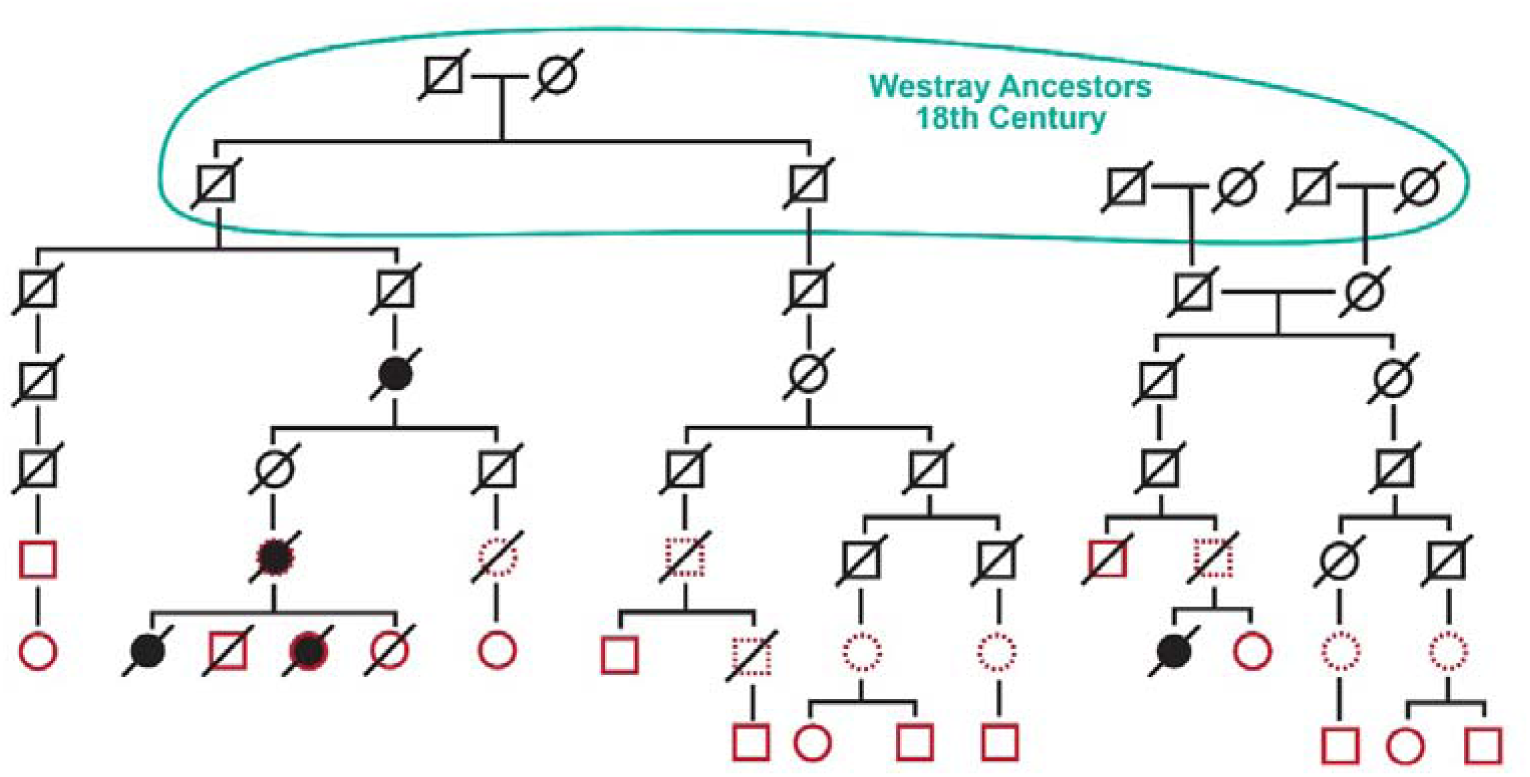
Outline pedigree of two kindreds from the ORCADES study. Filled circles are breast or ovarian cancers, red outlines are sequenced V1736A carriers, dotted red outlines are obligate carriers.

Consideration of causes of death from Scottish death registration certificates after 1855 reveals that the great-grandmother of four of the ORCADES carriers (and their close relative who died of breast cancer in her late fifties) died of breast cancer in the mid-1930s.

### Population frequencies of the BRCA1 *variant*

No other *BRCA1* variants reported as pathogenic or likely pathogenic in ClinVar were present in the ORCADES exome dataset, including the Scottish pathogenic founder mutation 2800delAA (p.Lys894fs)^30^. As would be expected, a number of variants of unknown significance in *BRCA1* were observed in this dataset. Information on allele frequencies in populations can be obtained from the Genome Aggregation Database (gnomAD)^31^. However, gnomAD v2.1.1, containing 125,748 exomes and 15,708 genomes from unrelated individuals sequenced as part of various disease-specific and population genetic studies, has no instances of the c.5207T>C; p.Val1736Ala variant, emphasising its rarity. Consistent with this, the variant is not observed at significant frequency in several cosmopolitan population research cohorts (Table 1). In the Viking Health Study Shetland (>2000 Shetlanders, who are genetically closest to Orcadians), no participants were observed with this mutation. The DiscovEHR browser also indicates no V1736A carriers were recorded in 92,453 individuals in the Pennsylvanian MyCode population cohort^32^. In contrast, the first 200,000 exome sequences from the UK Biobank^14,33^ contain four instances. This corresponds to a UK allele frequency of 0.00001, ∼480-fold lower than we found in Orkney. None of the four UK Biobank subjects was born in the Northern Isles; two live in Scotland and two in England. Three of the four UK Biobank research participant V1736A carriers are female. One, aged in her late fifties at assessment, has ovarian and fallopian tube ICD-10 cancer codes in the EHR dataset (Table 1). The two other female variant carriers, in their 50s and 60s at assessment, and the single male, have no reported ICD-10 codes relating to hereditary breast–ovarian cancer (HBOC). Small numbers of variant carriers are also reported in two databases of genomic data from cancer cases, CanVar and BCAC, the Breast Cancer Association Consortium (Table 1). However, the V1736A variant was not observed in sufficient numbers of cases and controls to allow for estimation of cancer risks in BCAC^34^.

### All V1736A carriers share a common haplotype

We were interested in obtaining DNA from the case or other members of the index family described in Domchek *et al*.^28^, to compare the haplotype background with that in Orcadian carriers, but no DNA was available.

All twenty heterozygous V1736A carriers in ORCADES share the same haplotype around the mutation, with a minimum length of ∼2 Mb (Figure 3). Access was given to exome sequence data surrounding the same mutation in a breast cancer patient (Table 1) described by Grzymski *et al*.^22^, for comparative haplotype analysis. Analysis of exome data from this carrier participant in the Healthy Nevada Project^22^ revealed that there were no opposing homozygote genotypes versus an ORCADES carrier across 676 SNPs, totalling 407 kb, consistent with them sharing one haplotype identical-by-descent in this region. Analysis of haplotypes in the genotype data from the four UK Biobank participants carrying the variant showed that they all shared a ∼1.1 Mb long haplotype, which was identical to the Westray haplotype from ORCADES.

**Figure 3.**
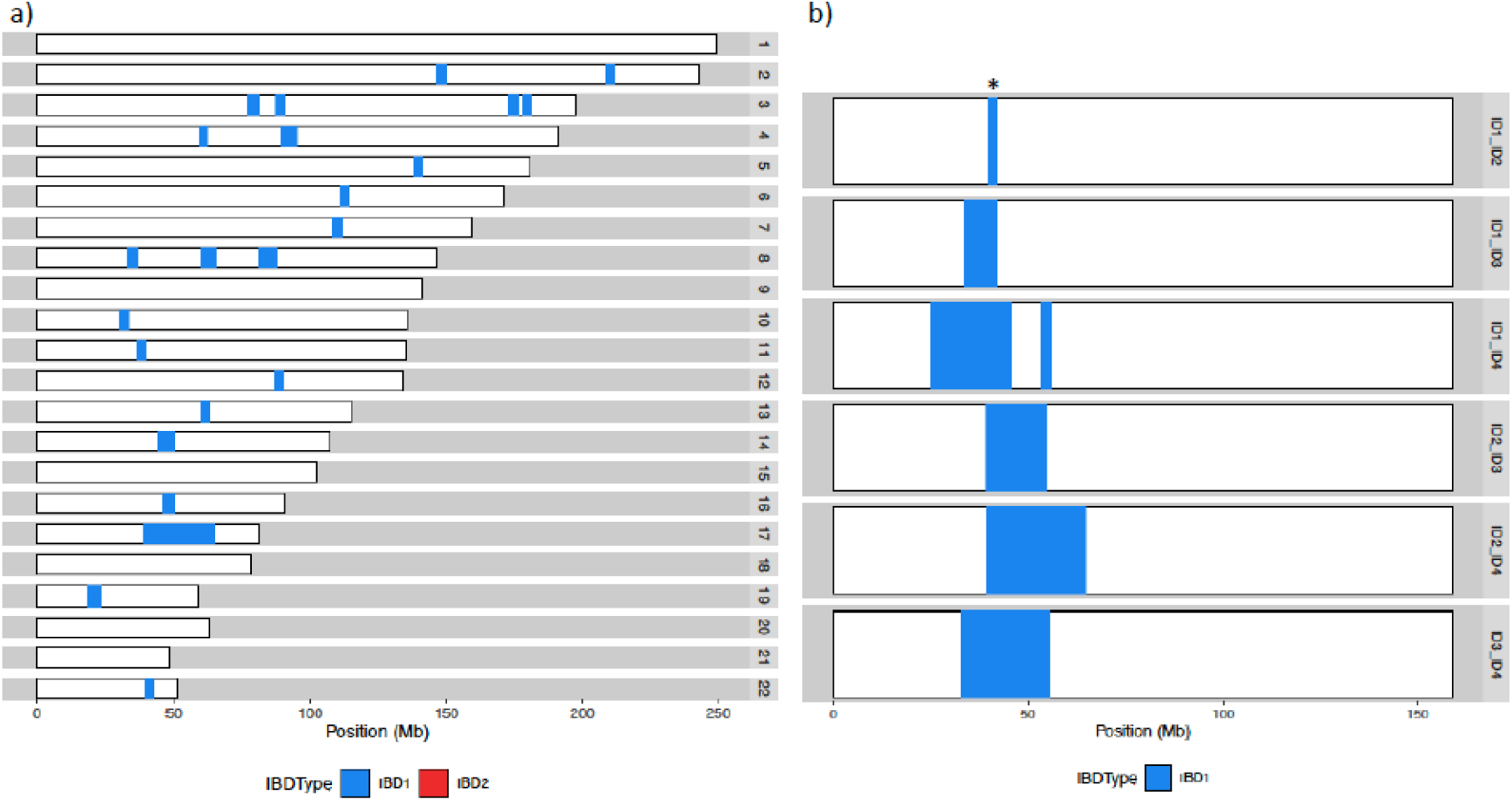
Haplotype sharing. (a) Genome-wide identity-by-descent sharing between two ORCADES carriers from different kindreds. In addition to the shared *BRCA1* haplotype on chromosome 17, background sharing due to Westray ancestry can be seen across the genome. (b) Haplotype sharing across chromosome 17 for all pairwise combinations of representatives of each of the four kindreds in ORCADES. Mb, megabase; IBD, identity-by-descent; * denotes the shortest shared haplotype

### Penetrance of V1736A in Orcadians

In addition to the exome sequence information, and the detailed study data collected in the recruitment clinics, linkage to routine NHS data in the electronic health record (EHR) provides a longitudinal component to many research cohorts, including ORCADES and the UK Biobank. The morbidity (hospital admissions, SMR01) and cancer registry (SMR06) datasets are particularly useful for research on people living in Scotland. These data have been obtained for almost all participants in the ORCADES cohort. The mean age of the seven female carriers at time of recruitment to ORCADES (baseline) was 54, and six gave permission for EHR linkage. Two V1736A carrier participants died over 80 years of age, one of whom had ovarian cancer recorded as cause of death. None of the remaining carriers had a diagnosis of breast or ovarian cancer recorded, and four with EHR linkage survive (Table 2).

Nine female obligate carriers linking two branches of the ORCADES pedigrees together were also ascertained (Table 2). Two died of breast cancer, three died of other causes and three remained cancer-free when this family history data was collected in the research study questionnaire (for one there was no information).

Together with the clinically-ascertained cases, there are a total of 37 women of Orcadian heritage with the variant, only two of whom overlap between the clinically-ascertained cases and ORCADES/obligate carriers. Importantly, comparison of common ancestors between the clinically ascertained and population-based pedigrees demonstrated that it is likely that only two out of the seven female variant carriers in the ORCADES population cohort have already been offered genetic testing as part of the cascade testing of the index family.

None of the female carriers in ORCADES are compound heterozygotes for pathogenic or likely pathogenic exonic *BRCA1* alleles, for which there are multiple submitters and no conflicts in ClinVar. Neither do they carry known pathogenic exonic variants in the cancer susceptibility genes *APC, BRCA2, RET, PALB2, MAX, TMEM127, BMPR1A, SMAD4, TP53, MLH1, MSH2, MSH6, PMS2, MEN1, MUTYH, NF2, SDHD, SDHAF2, SDHC, SDHB, PTEN, RB1, VHL* or *WT1*. One of the males carries a pathogenic *MUTYH* variant and one carries a likely pathogenic *PMS2* variant; none carry any other actionable variants in the listed genes.

## Discussion

Our combined approach of a family-based case study and systematic analysis within a population cohort has identified the pathogenic p.Val1736Ala *BRCA1* mutation in 1% of Orcadians. We have discovered a shared rare long haplotype background, which demonstrates the underlying relatedness of all carriers. The variant is likely to have arisen in a founder individual from Westray in the Orkney Islands in Scotland, at least 250 years ago. Analysis of the mutation in the population cohort has identified individuals not ascertained by classical cascade testing of relatives from the clinical pedigree. In a previous instance of a rare clinically relevant variant, p.Gly584Ser in *KCNH2* ^21^, we similarly identified carriers in our Northern Isles research populations who would not have been predicted from genealogy. Moreover, for both actionable variants, distant relatives were identified (e.g. seventh degree relatives) who could not be ascertained by cascade testing of first or second degree relatives. This highlights the value of research cohorts in informing clinical practice by more fully describing the burden of rare clinical variants segregating in these populations, which may in turn help planning and delivery of genetic services.

Participants of both clinical research and biobanking studies often wish to receive their results, particularly about “actionable” findings. This applies when they are asked to make both hypothetical and real decisions, and has recently stimulated the publication of an international policy on returning genomic research results^38^. However, the informed consent for most research cohorts, including the UK Biobank, does not allow the return of results about carrier status of actionable variants to individuals or their healthcare providers^14^. In contrast, we have begun a process of implementing the option of consent for return of selected clinically actionable results to consenting Viking Genes participants. Others also recommend the return of results following detection of hereditary breast and ovarian cancer risk to adult population-based biobank participants^38-40^. This genomic sequence-based approach permits the detection of individuals at high risk who would not be identified by personal and family disease histories alone. All results to be returned would first be verified in the research laboratory by an independent method, e.g. Sanger sequencing. Once the participant entered the clinical care pathway, a new blood sample would be taken for clinically accredited analyses by the NHS.

.Mutations in actionable genes like *BRCA1* are demonstrably estimated to be more penetrant in the clinical context of a family history of the relevant condition than in population-based cohorts. This is due in part to co-inheritance of multiple lower penetrance modifiers, and to ascertainment bias^41^. In the clinical genetics setting, *BRCA1* penetrance to 80 years of 79.5% (95%CI 75.5–83.5%) for breast cancer and 65% (95%CI 75–84%) for ovarian cancer were reported^42^, whereas population cohorts indicate lower risks^41,43^ Penetrance of pathogenic/loss-of-function variants in *BRCA1* was heterogeneous in population cohorts (mean [range], 38% [0%-100%])^41^. Despite the large kindred size we report here, power is limited to estimate precisely the penetrance of V1736A. The available data suggest more modest breast cancer penetrance than is typically seen in genetic clinic families with *BRCA1*, which fits with a missense variant with some residual function. However, the penetrance data we present is similar to that of many other pathogenic variants in *BRCA1*^42 44^.

Breast cancer risk in women from breast-ovarian cancer families born before 1940 is considerably less than in those born after^42^. This is likely not only due to reduced longevity, but also to lower body mass, higher parity, prolonged breast feeding and dietary factors in earlier generations. This observation fits with our data (available on request) that indicate higher ovarian than breast cancer risk. Risk-reducing bilateral salpingo-oopherectomy has been a common choice amongst women with positive predictive tests who have completed child bearing. Counselling in the family to date has highlighted the familial context. The majority of female gene carriers have accepted the offer of breast MRI screening, but use of risk reducing mastectomy has been limited. Recruitment of thousands of further research volunteers of Orcadian heritage in VIKING II^45^ will add more statistical power, through the discovery of new carriers and obligate carriers. VIKING II recruitment has highlighted scientifically for the first time the extent of Orcadian emigration, across Canada, New Zealand, and Australia but also in England and mainland Scotland. It is not possible to estimate accurately the size of the Orcadian diaspora, but it probably numbers tens of thousands of people.

High penetrance *BRCA1* and *BRCA2* founder mutations are described in a number of populations such as Iceland, the Ashkenazim, Poland, Norway and others, and testing for these is well established^5,6,8,9^. For example, around 1 in 40 Jews carry one of three BRCA1/2 founder mutations. In the French-Canadian founder population, twenty variants in *BRCA1, BRCA2*, and *PALB2* that predispose families to breast and ovarian cancer have been identified at increased frequencies. A recent paper demonstrated that genetic screening in that population could identify up to 10% of those who currently present with early-onset breast and ovarian cancer, prior to a diagnosis^46^. However, the challenges of likely reduced penetrance in those without a known family history of cancer, and cost, have limited adoption of asymptomatic BRCA screening outwith selected founder populations in resource-limited healthcare systems. The carrier frequency of 1% that we observe for the c.5207T>C; p.Val1736Ala *BRCA1* variant in the Orkney population is higher than some of the founder variants reported in these populations, and cost effectiveness of population-based screening for *BRCA1* founder mutation at 1% frequency has been reported in Sephardi Jewish women^10^. Very recently, NHS England announced its first programme of targeted founder BRCA mutation screening for people with at least one Jewish grandparent, where the population frequency of three pathogenic variants is around 1 in 40 (<u>NHS to launch expanded BRCA genetic testing for Jewish community - The Jewish Chronicle</u> (thejc.com)).

Our results suggest that the case should be made for carrier screening in the Orkney population for specific genetically drifted variants. We recommend that all women of Orcadian ancestry (worldwide) with a diagnosis of breast cancer should be offered a targeted test for this variant, if a BRCA1 & 2 gene screen is not offered as part of their clinical care. This targeted test for Orcadians with a family history of breast or ovarian cancer is now routine practice in the NHS Grampian clinic. However, expanding testing to all women of Orkney heritage, regardless of family history, would inevitably have resource implications for the NHS. Slightly over 11,000 females live in Orkney, of whom ∼9,300 are adult, and ∼70% of residents have two or more Orcadian ancestors. This suggests that to date we have identified less than half of the resident Orcadian V1736A carriers. We are therefore preparing a business case for population-based screening for the variant through primary care community hubs in Orkney, using the inexpensive single fragment Sanger sequencing assay that is established in the NHS Grampian genomics laboratory. We propose initially to offer the opportunity to Westray residents of known Westray ancestry, in order to offer the test first to those at highest risk. In support of this approach, an economic evaluation of population-based *BRCA1*/*BRCA2* mutation testing across multiple countries and health systems has recently been published^47^. Population-wide carrier screening in higher-risk isolated populations may help to decrease morbidity and mortality.

The V1736A founder mutation is only one of many contributors to breast and ovarian cancer risk in Orcadians, and high penetrance genes contribute only a proportion of cancer genetic risk. Population attributable risk of common low penetrance variants identified through genome-wide association studies explains a further component. Polygenic risk scores (PRS) are being considered for enhancement of risk stratification, both in the general population and in BRCA1/2 carrier population^48^. Work is ongoing to assess the utility of PRS in Orcadians, and to determine if low penetrance breast cancer-associated SNPs are enhanced or reduced in this population. Genetic drift of common low penetrance variants may limit the portability of scores developed elsewhere.

In conclusion, we propose that women with two or more Orcadian grandparents should be offered testing for the V1736A variant, regardless of family history of breast or ovarian cancer. The analyses presented here of the *BRCA1* variant are relevant beyond the modern population of Orkney, both as an exemplar and due to emigration to elsewhere in the British Isles and the New World. Future research will explore further genetically drifted loci observed as part of clinical care in Orkney and Shetland in the Viking Genes research cohorts.

## Data Availability

There is neither Research Ethics Committee approval, nor consent from individual participants, to permit open release of the individual level research data underlying this study. The datasets generated and analysed during the current study are therefore not publicly available. Instead, the research data and/or DNA samples are available from on reasonable request, following approval by the Data Access Committee and in line with the consent given by participants.

## Data Availability Statement

Some information (e.g. age and nature of a diagnosis) could potentially make individuals identifiable, so has not been shown, or is presented in aggregate form. These data can be made available to legitimate researchers affiliated to an academic organisation through application to the corresponding author. There is neither Research Ethics Committee approval, nor consent from ORCADES participants, to permit open release of the individual level research data underlying this study. The datasets generated and analysed during the current study are therefore not publicly available. Instead, the research data and/or DNA samples are available from on reasonable request, following approval by the Data Access Committee and in line with the consent given by participants.

## Acknowledgements

The study team wish to thank staff from the NHS Grampian genetics team and the ORCADES Study for their contribution to these datasets. In particular, Barbara Gibbons for genetic counselling of family members, the NHS Grampian genomics laboratory team for finding and testing for the variant, and Laura Taylor of NHS Grampian and the Public Health Scotland genealogy team for assembling the clinical pedigree. ORCADES DNA extractions were performed at the Edinburgh Clinical Research Facility, University of Edinburgh. Sanger sequencing was performed by Camilla Drake and the technical services team at the MRC HGU. Emily Weiss and Reka Nagy assembled the ORCADES pedigree using records at the General Register Office and study information, building on earlier pedigree work by Ruth McQuillan and Jim Wilson^36^. We thank Thibaud Boutin for phasing the GSA chip data and Kiera Johnston for help with analysis of other cancer susceptibility genes. The data in the EHR was provided by patients, and collected by the NHS as part of their care and support. The authors acknowledge the support of the eDRIS Team (Public Health Scotland) for their involvement in obtaining approvals, provisioning and linking this data. We would also like to acknowledge the invaluable contributions of the research nurses in Orkney and the administrative team in Edinburgh. Finally and most importantly, we thank the people of Orkney for their involvement in and ongoing support for our research.

## Author Contribution Statement

SK managed the project and drafted the manuscript. EC analysed the clinical data. LK analysed the exome datasets and did the haplotype comparisons. CB and DO’S recognised and interpreted the variant and provided clinical expertise. DB managed and analysed the EHR data. JJG contributed data. RGC performed the exome sequencing. CVvH, GT and ARS conceived and managed the ORCADES exome sequencing. JFW is the Chief Investigator of ORCADES, was awarded funding to implement the work, did genealogical analyses and interpreted the data. ZM recognised the family, initiated the work, led the clinical team, interpreted the data and proposed policy. All authors provided input and feedback on drafts of the manuscript.

## Ethical Approval

It is clear that information robustly linking genetic variants (e.g. *BRCA1*\*V1736A) with specific conditions (e.g. breast and ovarian cancer) is fundamental biological knowledge, not personal information, and therefore should not require specific consent for clinicians to share^49^. In contrast, neither identifiable medical details about the patients, nor their personal identifiers, were shared by the clinical team with the research team. Eligible participants were recruited to ORCADES, Research Ethics Committee references 26-11-2003 and 12/SS/0151. Research participants gave written informed consent for research procedures including electronic health record linkage. The data linkage and access to NHS Scotland-originated data for the ORCADES cohort was approved by the Public Benefit and Privacy Panel for Health and Social Care (Ref 1718-0380). This research has also been conducted using data from UK Biobank, as part of project ID number 19655.

For the purpose of open access, the author has applied a Creative Commons Attribution (CC BY) licence to any Author Accepted Manuscript version arising from this submission.

